# A Genome-wide Association study of Buccal Mucosa Cancer in India and Multi-ancestry Meta-analysis Identifies Novel Risk Loci and Gene-environment Interactions

**DOI:** 10.1101/2025.04.16.25325815

**Authors:** Sharayu Mhatre, Diptavo Dutta, Anand Iyer, Shruti Gholap, Aseem Mishra, Manigreeva Krishnatreya, Grace Sarah George, Pravin Narayanrao Doibale, Yuzheng Dun, Ziqiao Wang, Om Jahagirdar, Pankaj Chaturvedi, Preetha Rajaraman, Cheng-Ping Wang, Anil Chaturvedi, Siddhatha Kar, Rajesh Dikshit, Nilanjan Chatterjee

## Abstract

Genome-wide association studies (GWAS) of oral cancers (OC) to date have focused predominantly on European Ancestry (EA) populations. India faces an excess burden of OC, but the most common site of occurrence is the cancer of the buccal mucosa, which is relatively rare in EA populations. We conducted a GWAS of buccal mucosa cancer (BMC) comprising 2,160 BMC cases and 2,325 controls from different geographical locations in India. Single-SNP association tests detected one novel locus (6q27) and one novel signal within the known OC risk locus 5p13.33, at the genome-wide significance level (P-value<5X10^-8^). We additionally conducted a GWAS of 397 BMC cases and 439 controls from Taiwan and performed multi-ancestry GWAS meta-analysis of OC on 5255 cases and 8748 controls across EA, Indian and Taiwanese populations. We identified a novel risk locus harbouring the tumour suppressor gene *NOTCH1* through a gene-level analysis of the multi-ancestry GWAS data. Pathway analysis suggested that PD-1 signalling, and Interferon Gamma Signalling may be important in the aetiology of BMC. Within data from the Indian BMC GWAS, we further identified statistically significant evidence of both multiplicative interactions (P-value=0.026) indicating stronger polygenic risk of BMC among individuals with history of chewing tobacco compared to those without. Our study provides insights into the etiologies of BMC in India, highlighting both its similarities and differences with other types of oral cavity cancers, as well as the interactions between polygenic gene score and tobacco chewing.

## Introduction

Head and neck cancers(HNC), a broad category that includes cancers of the oral cavity(OC), pharynx and larynx, are a common form of cancer globally, affecting 892,128 new individuals annually worldwide with about 37% of cases arising from South and South-East Asia alone. ^1^ There are geographical differences among subsites of HNC with occurrences arising in the tongue, lips and oropharynx being more common in European and American populations while cancers of the buccal mucosa and alveolus are predominant in South East Asia and India. In India, Buccal Mucosa Cancer (BMC) is the most common cancer site among males with 113,000 new cases arising annually. ^2,3^ Common risk factors for head and neck cancers in European populations include tobacco smoking, alcohol drinking, and infection with Human Papilloma Viruses (HPV). ^4^ In contrast, for Indian and some Asian populations, the use of smokeless tobacco or chewing tobacco is considered the most important contributor of risk. There is limited understanding of the role of HPV infection in OC cancers in these Asian populations. ^5^

Genome-wide association studies(GWAS) mainly conducted in European ancestry populations have identified to date 11 susceptibility loci for OC and oropharyngeal (OPC) cancers together. ^6–9^ A prominent finding has been the identification of the Human Leukocyte Antigen (HLA) region as a risk locus in European and North American populations where HPV infection has been attributed as a major cause. This result was not replicated in the Latin American study where HPV-associated OC/OPC is rare. ^9^ Other prominent findings include alcohol associated genetic loci *ADH1B-ADH7* (4q23), and multi-cancer risk loci *TERT-CLPTML* (5p15.33) and *CDK2NA-CDK2NB* (9p21.3). ^9^ While India faces a major burden of OC, there has been no large-scale GWAS to date in this population. Given the uniqueness of the predominating site (BMC) with tobacco chewing as a major risk factor, it remains unknown whether the genetic etiology of OC in India and other Asian countries overlaps with those from other continents.

We conducted the first large-scale GWAS of BMC risk using 2,160 cases and 2,325 controls arising from different regions of India. We further conducted additional GWAS using 397 BMC cases and 439 controls from Taiwan and carried out a genome-wide multi-ancestry meta-analysis of GWAS data across India, Taiwan, Europe and North America and South America consisting of a total of 5,123 OC cases and 8,558 controls. Altogether, our analyses identified two novel loci and multiple novel variants within known loci that provide distinctly better representation of risks in the Indian population than previously identified variants from prior studies. We further carried out a series of post-GWAS *in silico* analyses, including multi-tissue transcriptome-wide association studies (TWAS), heritability estimation and cross-population genetic correlation analysis, and Summary-data-based Mendelian Randomization (SMR) analysis to obtain deeper insight into the genetic architecture of BMC and its overlap with other forms of OC. Finally, using a polygenic score (PGS) for five identified risk variants for the Indian population, we explored the nature of gene-environment interactions with respect to chewing tobacco habits leading to unique insights into the nature of genetic effects in the presence of an exceptionally strong environmental risk factor.

## Material and Methods

In this case-control GWAS, we did the GWAS discovery study on 2,160 buccal mucosa cancer (BMC) cases and 2,325 hospital visitor controls coming from hospitals of Tata Memorial Centre including Tata Memorial Hospital (TMH), Mumbai; The Advanced Centre for Treatment, Research and Education in Cancer (ACTREC); Nargis Dutta Memorial Cancer Hospital (NDMCH); Barshi, Bhubaneswar Borooah Cancer Institute (BBCI), Guwahati; Homi Bhabha Cancer Hospital (HBCH) and Mahamana Pandit Madan Mohan Malviya Cancer Centre (MPMMCC), Varanasi (Supplementary table 1 & Supplementary figure 1).

Cases were patients with buccal mucosa cancer (International classification of diseases for Oncology Version 3[ICD-O-3] site code Upper alveolus (C3.0), lower alveolus (C03.1), Buccal Mucosa (C06.0), Retromolar areas (C06.2), Bucco alveolar sulci, upper and lower (vestibule of mouth) (C06.1)) (Supplementary table 2) visiting Tata Memorial center for treatment and diagnosis in between October 10, 2010 and September 29, 2021.

Cases eligibility criteria were primary BMC confirmation on basis of histopathological or cytological diagnosis from buccal mucosa; a date of diagnosis of less than one year before the date of enrolment, age between 19 years and 75 years; and being resident of India for at least 1 year. We excluded the patients from any other malignancies of HNC, with clinical diagnosis of BMC but without microscopic confirmation was excluded. Institutional electronic medical record (EMR) or diagnostic report was used to obtained histopathological/ cytological confirmation of BMC cases ascertainment.

We recruited hospital visitor controls aged 19-75 years with no history of cancer visiting all departments or units of Tata Memorial Centres in similar geographical locations as to cases who had been residents of India for at least 1 year and frequency matched them to cases on the basis of age, sex, and current region of residence. Controls were recruited from wards, waiting areas and outpatient departments of eleven diseases management groups (DMG) including adult hematolymphoid, thoracic, urology, bone-soft tissue, breast, gastrointestinal, gynecological, HNC services, neuro oncology, pediatric hematology, pediatric solid tumours and department of preventative oncology. We have not recruited more than 32% of controls from any of the DMGs. The controls recruited from various DMG ranges from 1 to 32% (mean number of controls recruited from each DMG =7.69%) (supplementary table 3). The first-degree controls were not recruited from head and neck cancer DMG. We allowed comorbidities for both cases and controls. The controls were recruited after group matching for age, gender and geographical region.

The following data were collected by a standardised interviewer administered questionnaire for all study participants at the time of enrolment: demographic and socioeconomic information, dietary information, use of alcohol and tobacco, detailed residential history, reproductive history, and personal and family medical history. We recruited cases and controls simultaneously during the study period.

To conduct multi-ancestry meta-analysis, we genotyped 397 BMC cases and 439 controls enrolled over a period of 3 years for a Natural History of Oral Cancer Precursor Lesions: A Prospective Cohort Study conducted in the Taiwanese population. We obtained the informed consent from all study participants and study was approved by respective institutional ethics committee (Supplementary table 4).

### Genotyping

We extracted genomic DNA from all peripheral blood samples using the Qiagen QiAmp® Blood DNA Midi Kit (spin-column protocol) and/or QIAsymphony® DNA Midi Kit (paramagnetic bead protocol), Qiagen, Hilden, Germany. The DNA was quantified using the Picogreen dsDNA Assay kit (Invitrogen, Carlsbad, CA, USA) and diluted to a concentration of 50 ng/µL. We genotyped GWAS study samples on the Infinium Global Screening Array version 3.0 Illumina Bead Chip array harbouring ∼654,027 markers (Illumina, San Diego, CA, USA. The cases and controls were genotyped simultaneously, following 1:1 or 1:2 (cases: controls) configuration. At the minimum, one inter-assay and one intra-assay were processed in 96 samples. We collected the scanned intensity data and clustered and called genotypes (i.e., identified genotypes) using Illumina Genome Studio version 2.0. On the basis of the GenTrain3 calling algorithm, we estimated genotype clusters from samples with a preliminary completion of greater than 95% over SNPs. We proceeded with 151 expected experimental duplicates of which 90 were inter-assay and 61 were intra-assay samples. All laboratory procedures for processing samples from India and Taiwan were conducted at the Division of Molecular Epidemiology and Population Genomics at the Centre for Cancer Epidemiology, Tata Memorial Centre, located in Navi Mumbai, India.

### GWAS Quality Control - samples

We applied extensive quality control metrics to the genotyping data, of both the samples of Indian as well as Taiwanese origin, using PLINK 1.934. ^10,11^ We removed the individuals with a call rate of less than 95% (cut-off level empirically determined). We eliminated individuals with unresolved and reported sex differences. For males, we exclude those with more than 5% heterozygosity based on the X chromosome SNPs. For females, we exclude those with less than 15% heterozygosity based on X chromosome SNPs. Further, samples with unsolved genetic discrepancies, leading to cryptic relatedness between them, were identified by performing Identity-by-decent (IBD) analysis.

Samples with lower genotyping call rate from 123 expected experimental duplicate-pairs from the Indian nationals’ data (IBD >0.9) were excluded. In addition to this, we identified 57 unexpected duplicate-pair (IBD > 0.1875) and excluded one sample from each pair, prioritizing cases over controls and for pairs with the same status, we retained the older study participants. The specifics regarding the exclusion of quality control from the Indian data set are outlined in Supplementary table 5 and illustrated in Supplementary figure 1.

Similarly, for the Taiwanese population data, we removed 28 expected experimental duplicate-pairs from the Indian population data (IBD >0.9). Additionally, we identified 2 unexpected duplicate-pair (IBD > 0.1875) and excluded one sample from each pair.

To address the underlying population structure, we did principal components analysis(PCA) for the Indian data set using version 7 of Eigensoft’s SmartPCA software, for identifying underlying population strata, if any. ^12,13^ We excluded outliers from the population in the discovery set (n=142) and the Taiwan replication set (n=21) to obtain eigenvectors for adjusting the GWAS analysis The Principal Component Analysis (PCA) of each individual for the Indian population can be found in (Supplementary figure 2). Finally, GWAS included 4199 individuals (2046 cases and 2153 controls) from Indian population and 800 individuals (379 cases and 421 controls) from the Taiwan population.

### SNP quality control and imputation

At the start, the Indian as well as the Taiwanese data set had 654027 variants. SNPs with a call rate of less than 95% (cut-off level empirically determined) were excluded, resulting in a removal of 5765 variants in the Indian data & 8901 variants in the Taiwan data set respectively (Supplementary table 6). Markers with a failure to meet Hardy-Weinberg equilibrium exact test at a P-value of less than 10^-6^ were excluded, resulting in a removal of the 1051 variants in the discovery & 84 variants in the replication data set respectively. Variants with a minor allele frequency (MAF) of less than 0·05 were excluded from the genotyped data, resulting in a removal of 326,668 variants in the discovery & 361,162 variants in the replication data set respectively (Supplementary table 6).

For PCA, a further cleaning of any duplicate or multi-allelic SNPs was done. A total of 536 duplicate variants from the Indian data set & 439 from the Taiwanese data set were removed. 95 variants with allele codes outside of A, C, G & T from the discovery set & 75 such variants from the replication set were removed. PCA was done on 319,912 & 283,366 variants for the Indian and Taiwanese data set respectively.

### Imputation

After cleaning the SNPs data, we filtered out variants with MAF < 0.05 and call rate < 0.05. As a result, we were left with 320,543 SNPs in the Indian dataset and 283,366 SNPs in the Taiwanese dataset, which were suitable for imputation. We phased the genotypes using Eagle software and then imputed them using the Michigan imputation server [version 1 using Minimac 4 & Eagle 2 for phasing].^14–20^

We have utilized version 5 of phase 3 data from the 1000 Genomes Project as our reference set. Our genotype data was matched with the 1000 Genomes Project Phase 3 data by using Genotype Harmonizer v1.4.23.^21–24^ The output was formatted to use the same reference alleles as the reference data set, while also preserving the unique variants present in our genotyped data. We received 47,099,450 variants from the Michigan server for the Indian data set. To ensure maximum variant coverage, we merged this data with our genotyped data. Then, we filtered out any duplicate position variants (removed 776,935 variants) and multiallelic variants (removed 3,110,762 variants). We retained only the variants with ACG and T alleles and removed any variants with an imputation r^2^ value below 30% (removed 29,023,210 variants). After these filtering steps, we were left with 14,509,086 variants for the Indian data set (Supplementary table 6). We applied a comparable approach to the Taiwanese dataset and filtered it using high-quality imputation statistics. This resulted in the retention of a total of 10,207,065 variants (Supplementary table 6).

Additionally, variants with a MAF lower than 0.5% were excluded from both datasets, resulting in the removal of 5,795,812 and 2,983,651 variants from the Indian and Taiwanese data set, respectively (Supplementary table 6).

We also performed an HLA region specific imputation on Michigan imputation server using the Four Digit Multi-Ethnic HLA reference panel data set (Version 2) ^56–58^ on the same aligned chromosome 6 file used above. The resulting imputed data was merged with the genotyped SNPs data set to generate a separate, HLA specific file. The analysis results from this data set also gave similar results to the main data set with imputation done across the genome, with similar top SNPs appearing.

### GWAS and meta-analysis

After pre-processing, we were left with 4199 samples and 8,713,274 markers in the Indian data set, as well as 800 samples and 7,223,414 markers in the Taiwanese replication data set. We performed an association analysis for the SNPs using PLINK version 1.9. ^10,11^ We used multivariable unconditional logistic regression, assuming a log-additive genetic or dosage model, with gender, age, and the top 10 eigenvectors derived as principal components from Eigensoft’s Smart PCA as covariates. We identified independent lead/index/sentinel variants using the LD-based clumping algorithm implemented in the commonly used post-GWAS analytic pipeline, FUMA (version 1.4.2), with the 1000 Genomes phase 3 version 5 South Asian populations as reference, applying a *r*^2^ threshold of 0.05 and a P-value threshold for genome-wide significance of P-value<5 x 10^-8^ to variants with minor allele frequency > 1% and imputation quality > 0.3 and considered lead variants within 500 kb of each other to be part of the same locus. ^21,22,28^

We performed a fixed-effect meta-analysis using the METAL tool to combine data from the India and Taiwan BMC GWAS and the Europe, North and South America OC GWAS. ^29,30^ We included summary GWAS statistics downloaded from the IEU Open GWAS Project server (three IDs, namely ie-b-93, ieu-b-94, and ieu-b-95) for the OC GWAS by Lesseur et al. ^31–33^ Additionally, we incorporated the Taiwanese GWAS in the meta-analysis. We used the classical approach of using effect size estimates and standard errors, which weights effect size estimates using the inverse of the corresponding standard errors. We performed METAL heterogeneity analysis using the I2 statistic to account for heterogeneity and determine whether observed effect sizes (or test statistics) are homogeneous across samples. We applied genomic control correction to all input data files considering the vast differences in geographical regions. Meta-analysis was completed on 5,032,849 markers in common to all five datasets (Supplementary table 6).

### Bayesian false discovery Probability (BFDP)

Apart from using the frequentist threshold of P-value<5×10^-8^, we also used approximate Bayes factors to compute the BFDP.^34,35^ BFDPs were calculated for each independent, genome-wide significant lead variant based on a prior probability of true association at any variant of 1 in 10,000 and assuming a plausible prior distribution for the log of the odds ratio (effect size estimate for a truly associated variant) to be a normal distribution with mean zero and standard error 0.2 (reflecting an odds ratio of 1.2). All analyses in this study were conducted using R version 4.2.1 unless otherwise specified.

### Heritability analysis

Genome-wide complex trait analysis (GCTA version 1.94) was used to estimate heritability of GWAS data which uses the “Genome-based Restricted Maximum Likelihood – LD Score” (GREML-LDMS) method for estimation while correcting for the LD bias in the estimated SNP-based heritability. ^36^ From a total of 47,419,993 variants that were extracted, we computed genetic relatedness matrix (GRM) using 24,537,203 variants. Subsequently heritability was estimated in the liability scale using the case-control status of each sample in the data and assuming a prevalence of BMC in the population to be 0.2%.

### Conditional analysis

We performed conditional analysis within associated regions using GCTA-COJO. ^37^ In this analysis, we used the summary-level statistics from Indian data set GWAS as input. For each locus, we used a window of +/- 10Mb to search for conditionally independent associations, among variants with a minor allele frequency (MAF) greater than 0.005, assuming complete LD between SNPs that are more than 10Mb away from each other.

### Identification of genetic risk loci and functional annotation

We identified independent lead/index/sentinel variants using the LD-based clumping algorithm implemented in the commonly used post-GWAS analytic pipeline, FUMA (version 1.4.2), with the 1000 Genomes phase 3 version 5 South Asian populations as reference, applying a *r*^2^ threshold of 0.05 and a P-value threshold for genome-wide significance of P-value<5 x 10^-8^ to variants with minor allele frequency > 1% and imputation quality > 0.3 and considered lead variants within 500 kb of each other to be part of the same locus. ^28^ We annotated lead variants with their most likely target genes based on the Open Targets online platform that integrates gene expression, circulating protein levels, chromatin conformation and interaction, and functional genomic profiles from multiple tissues and cell types to associate variants to genes.

### Gene and pathway level analyses

We did gene-level association testing for genes across the genome using the “multi-marker analysis of genomic annotation” (MAGMA) software version 1.08 implemented in the FUMA pipeline version 1.4.2.^28,38^ MAGMA involves mapping variants to the genes that they physically overlap (0 kb window size), accounting for LD between these variants, and doing statistical multi-marker association tests. We also estimated potential causal effects of gene expression on BMC risk by leveraging blood-based *cis*-regulated gene expression quantitative trait locus (eQTL) data for 31,684 European-ancestry individuals curated by the eQTLGen consortium.^39,40^ We performed “summary-data-based Mendelian randomisation” (SMR) coupled with the “heterogeneity in dependent instruments” (HEIDI) and tested for colocalization to nominate specific genes whose expression in whole blood was associated with cancer risk along with a clear evidence of overlap of the GWAS and eQTL association signals.^39^ MAGMA and SMR were applied to genome-wide association testing results from the Indian GWAS and the multi-ancestry meta-analysis. The genome-wide significance threshold for the gene-level MAGMA analyses was set at P-value=2.6 x 10^−6^ and for the SMR analyses was set at P-value=3.3 x 10^−6^ to account for testing 18,802 and 15,174 genes, respectively. For SMR, only genes with SMR p<D3.3 x 10^−6^ that also had a HEIDI test p>0.05 were declared genome-wide significant since p_HEIDI_>0.05 indicates colocalization of the GWAS and eQTL signals for a given gene and reduces the likelihood of confounding of the transcriptomic association by genetic linkage.

All genes with p<0.05 from the application of MAGMA to the Indian GWAS results were used as input for pathway analysis. Pathway analysis was done using the Enrichr online tool that involves application of the Fisher’s exact test to calculate pathway-level p values followed by adjustment for multiple comparisons using false discovery rate control by the Benjamini-Hochberg method. ^41–44^ Biological pathways were drawn from the 2022 version of the Biocarta database. ^45–47^

### Transcriptome-wide association study

We performed transcriptome-wide association studies (TWAS) using prebuilt expression imputation models for 6 tissues profiled in GTEx v8. In particular, we used data from five tissues in the upper gastrointestinal tract (Esophagus: gastroesophageal junction, Esophagus: Mucosa, Esophagus: Muscularis, Lung, Minor Salivary Gland) and Whole Blood. Tissue-wise false discovery rate (FDR) was estimated to adjust for multiple testing, and we declared any gene with FDR < 5% to be significant.

### Fine-mapping

We performed fine-mapping of the 5 independent signals identified in Indian BMC GWAS. For each signal, we retained variants in the Indian population with MAF ≥5% within a region 1DMb upstream and downstream of the sentinel (index) variant and extracted in sample LD across 4,199 samples from Indian population. Subsequently, using the GWAS summary statistics and the extracted LD estimates, fine-mapping was performed using sum of single effects (SuSiE; v0.12.35). ^48,49^ SuSiE performs a Bayesian stepwise regression at each locus to identify (A) the optimal number of potentially causal signals and (B) the minimal set of variants that cumulatively encompasses a pre-specified posterior probability (95%) of being the causal variant, called the credible set.

### Exposure information on use of Tobacco and Alcohol

Self-reported information on tobacco smoking, chewing habits, and alcohol use habits of the study participants was collected by trained interviewers using a pre-tested questionnaire.^50,51^

Tobacco smoking: An individual was considered a regular smoker if they smoked any form of tobacco at least once a week for a period of six months. Details of 11 smoking products, namely – cigarette, bidi, cheroot, cigar, roll you own, chutta, reverse chutta, dhuranti, reverse dhuranti, hookah, and hookli/chillum – were collected for the age of participant when they started using the product, age when they stopped, number per day, and how many days in a week they consumed it.

Tobacco Chewing: History of chewing tobacco consumption was collected for a wide variety of region-specific tobacco products. To assess lifetime use, the participants were asked whether they had ever chewed tobacco. (ever or never), either currently or in the past. An ‘ever chewer’ was defined as an individual who chewed tobacco at least once a week for a minimum duration of six months. Additionally, for a particular product, information such as the age at initiation, age at cessation, the quantity consumed (number per day) and the duration of consumption (in a week) were also collected.

### Alcohol use

A regular alcohol user was defined as consuming any alcoholic beverage at least once a month for a period of six months. We collected information on six different internationally-recognised liquors, and also a wide range of Indian region-specific locally-brewed liquor types. The participant’s age when they began consuming the liquor, and the age when they stopped consuming were recorded. The number of times the beverage was consumed in a day, the number of days in a week, and the quantity consumed as either millilitres (ml), glasses, shots, cans, or bottles, was collected as well.

Regular smokers, chewers of any tobacco, and regular consumers of any alcoholic beverage were analysed as ever-users versus their respective never users.

## Results

We enrolled 2,160 buccal mucosa cancer cases and 2,325 hospital visitor controls at the Tata Memorial Hospital (TMH), Mumbai; Advanced Centre for Treatment Research and Education in Cancer (ACTREC), Navi Mumbai; Nargis Dutt Memorial Cancer Hospital (NDMCH), Barshi; B Borooah Cancer Institute (BBCI), Guwahati; Homi Bhabha Cancer Hospital (HBCH) and Mahamana Pandit Madan Mohan Malaviya Cancer Centre (MPMMCC), Varanasi. Most of the cases (75%) and controls (78%) were enrolled at TMH, Mumbai which is followed by HBCH and MPMMCC, Varanasi and BBCI, Guwahati. (Supplementary table 1 & Supplemental figure 1). For cases, the most frequent ICD-O-3 code was buccal mucosa (C06.0; 73.56%) while the histopathological type of nearly all cases was squamous cell carcinoma (98.89%) (Supplementary table 2 & 7). In multi-ancestry meta-analysis, we further included a total of 397 BMC cases and 439 controls from Taiwan as well as summary-statistics data from a GWAS based on 2,698 oral cavity cancer cases and 5,984 controls of predominantly European ancestry. ^9^ After quality control checks, we conducted final analysis based on 2,046 cases and 2,153 controls on Indian population and 5,123 cases and 8,558 controls in multi-ancestry meta-analysis.

### Single SNP analysis

A total of 8,694,880 single nucleotide polymorphisms (SNPs) and indels with minor allele frequency of 0.05% or greater that had either been genotyped (assayed) or imputed with imputation quality score of 0.3 or greater were included in the GWAS of BMC in the Indian population (Supplementary table 6). GWAS results showed minimal evidence of genomic inflation (genomic control factor, λ_GC_=0.98; Figure 1). We identified independent signals at genome-wide significance (P-value<5×10^-8^) based on linkage disequilibrium (LD)-based clumping with an r^2^ threshold of 0.05 (Table 1). All the lead SNPs except one, identified at genome wide significance level had a Bayesian False discovery probability of less than or equal to 3% (Supplementary table 8). ^34,35^

**Figure 1:**
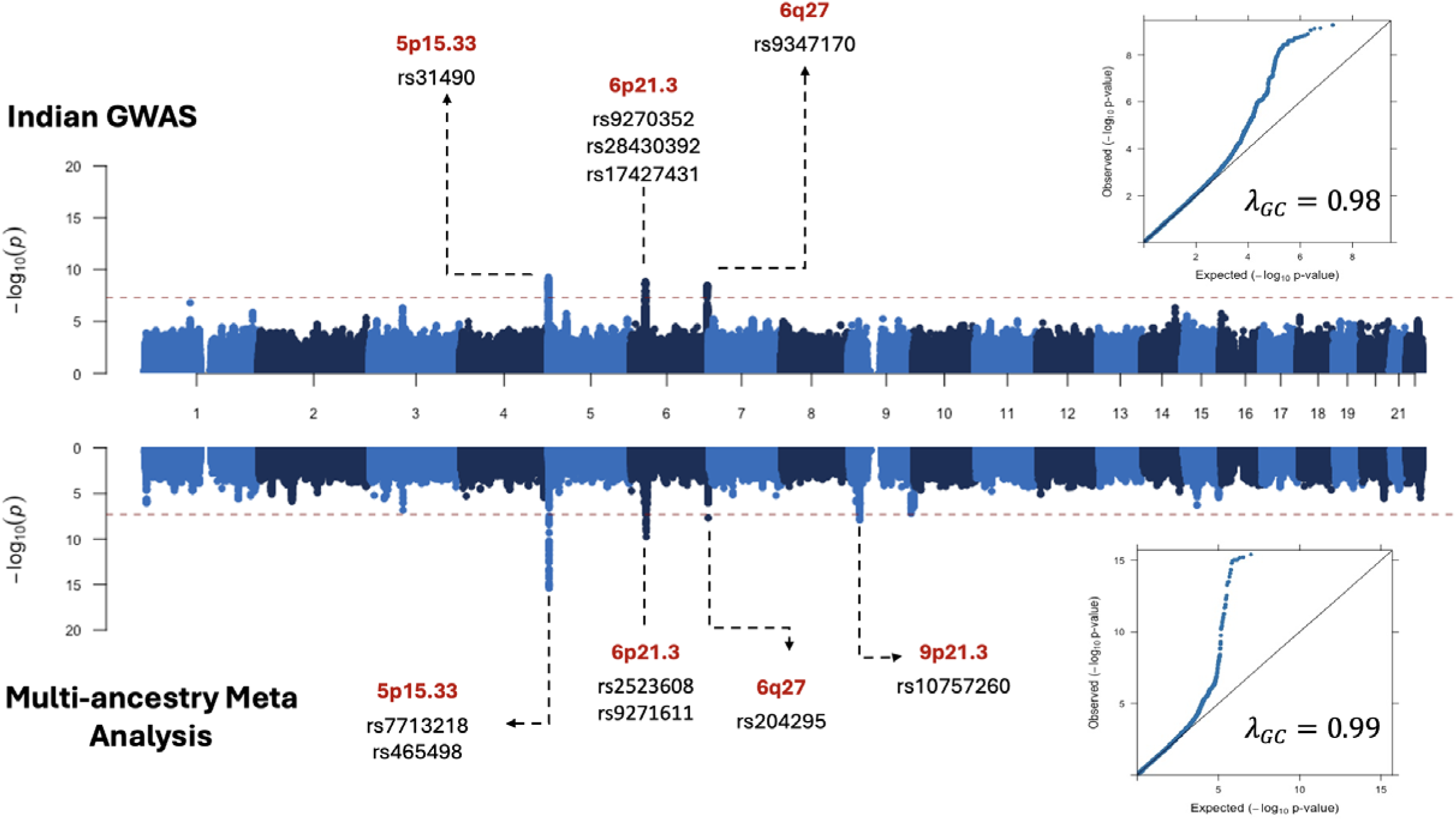
Genome-wide association study of BMC. The Miami Plot illustrating the Genome-wide significant SNPs (p < 5 x 10^-8) identified in multicentre study on Indian Buccal Mucosa Cancer. The lower panel illustrates a Genome-wide significant SNPS in multi-ancestry meta-analysis with Indian, Taiwanese, European population. Top and bottom inset shows the quantile–quantile (QQ) plot for the corresponding GWAS with the estimated genomic inflation factor.

**Table 1.**
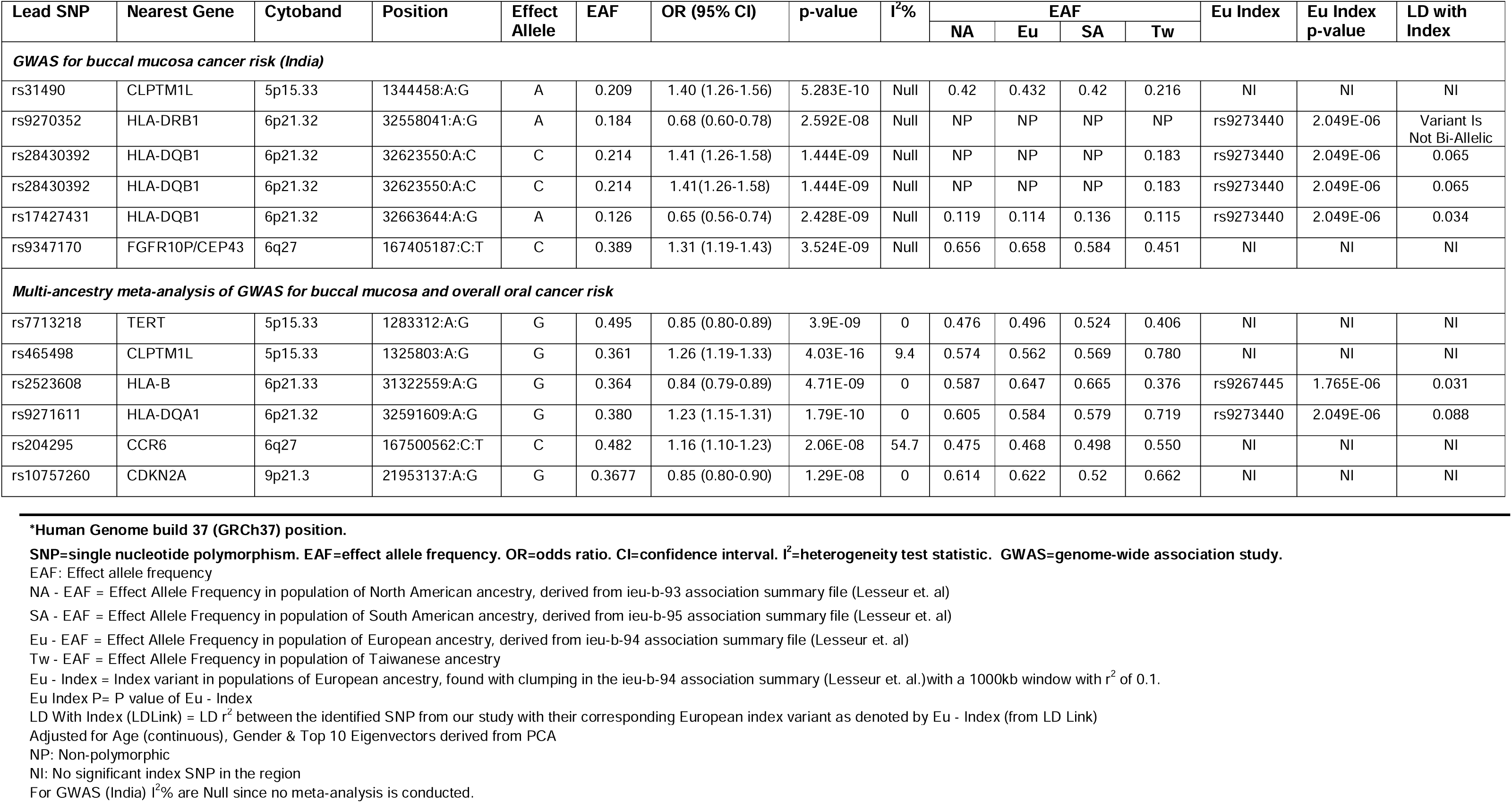
Genome-wide significant (p<5×10^-8^) independent (linkage disequilibrium r^2^ <0.05) lead SNPs identified in multi-center Indian study on Buccal Mucosa Cancer and in multi-ancestry meta-analysis on Indian, Taiwanese, European population.

Analysis based on the Indian BMC study alone identified five independent signals, four within known OC loci reported from prior studies (*5p15.33* and *6p21.32 HLA* regions) and one in a novel locus (6q27) at the genome-wide significance level. The strongest signal involved the lead SNP rs31490 at 5p15.33 (Table 1 and Figure 1) that was 756 bp from the canonical transcription start site (TSS) of *CLPTM1L* and was among 13 genome-wide significant oral cancer risk SNPs previously identified in individuals of European ancestry. ^52–54^ In an analysis within 5p15.33, but conditional on the lead SNP rs31490, we further detected a novel association with rs62332591 (OR=0.77 for G allele; p =1.84 x 10^-8^; 95% CI: 0.75-0.78; Supplementary table 9) which is in low linkage disequilibrium (LD) with rs31490 in the Indian population (r^2^=0.007, D^’^=0.1499) and is proximal to the well-known *TERT* gene in the region. ^55,56^ The second most significant locus, 6p21.32 (Table 1 and Figure 1), was marked by the lead SNP rs9270352 that was 416 bp from the canonical TSS of *HLA-DRB1*, which has been reported as a OC locus in prior studies. ^52–54^ Within the HLA region, we identified two other independent signals marked by lead SNPs rs28430392 and rs17427431, close to the *HLA-DQB1* gene (6p21.32). In a multivariate analysis, all three lead SNPs in the HLA region showed independent signals. Finally, the fifth signal involved the novel genomic region 6q27 (Table 1 and Figure 1), marked by the lead SNP rs9347170 (Table 1, Figure 1) with nearest gene being *FGFR1OP* (also known as *CEP43*), and has not yet been reported by any prior studies of OC or OPC in any population.

We next attempted to replicate all known OC risk loci reported in prior studies in Indian BMC GWAS (Supplementary table 10). Results showed that published lead SNPs in five out of seven risk loci reach nominal threshold P-value<0.05 in our study. Lead SNPs reported for genetic risk loci at chromosome 2p23.3 and 4q23.3 did not show any evidence of association with the risk of BMC in the Indian population. Out of 5 SNPs observed for OPC in earlier GWAS we could replicate one SNP (rs34518860) on risk loci 6p21.32 in Indian BMC GWAS (OR = 0.82, p =0.036). ^7^ The SNP rs1265081 on chromosome 6p21.33 observed for all head and neck cancer sites combined in earlier GWAS (effect allele C; OR 0.85, p =3.7×10-10) was significant in Indian BMC GWAS but in opposite direction (effect allele C, OR =1.12, P-value =0.01; Supplementary table 10). ^7^

In our multi-ancestry meta-analysis (Figure 1) of OC GWAS across India, Taiwan and Europe, we detected six independent signals: two in regions of *TERT-CLPTLML* (5p15.33), two in regions of HLA loci *HLA-B* (6q21.33) and *HLA DQA1* (6q21.32), one close to the gene *CCR6* (6q27) and one close to the gene *CDK2NA* (9p21.31). In 5p15.33, one of the two lead signals were close to the *CLPTML* gene, which has been associated with risk of OC in multiple prior studies. ^57–59^ The second lead signal in 5p15.33 indexed by rs7713218, is 11 kb from the canonical TSS of *TERT* (Figure 1 and Table 1), is a novel genome-wide significant association for risk of OC. The two independent HLA signals correspond to previously known associations in this region. There was no evidence of heterogeneity in effect sizes for the identified lead SNP in *TERT*. However, for *CLPTM1L* (5p13.33) and *CCR6* (6q27) regions, a heterogeneity of 9.4% (P-value = 0.35) and 54.7% (P-value = 0.07) was reported in the multi-ancestry analysis, albeit not significant.

The novel region 6q27 we identified based on the Indian study alone was also found to be genome-wide significant in the multi-ancestry meta-analysis (Table 1). However, the effect-size associated with the lead SNP (rs9347170) showed significant heterogeneity (P-value=1.61×10^-3^) by study and the SNP did not show any evidence of association in the large EUR-ancestry GWAS (P-value=0.25), indicating the signal in this locus is primarily driven by the Indian population (P-value=3.52 x 10^-9^).( Supplementary table 11). ^9^

Finally, the multi-ancestry meta-analysis identified the lead SNP rs10757260 in the known OC locus 9p21.3 near the gene *CDKN2A* (Table 1, Figure 1), but this new SNP is in weak linkage disequilibrium (LD) with previously reported lead SNP rs8181047 (Supplementary table 10); r^2^=0.09 in South Asian and r^2^=0.14 in European ancestry populations) for this region. ^55,56,60^ The original lead SNP rs8181047 showed evidence of only weak association (OR=1.13, P-value=0.025, for an effect allele=A) in the India data, while the new lead SNP showed stronger association (OR=1.19, P-value=1.74×10^-4^), and there was no statistical evidence of heterogeneity (I^2^=0) in effect size estimates across studies at this new lead SNP. (Table 1, Supplementary table 10 and 11)

### Gene-level Association Analysis

We further conducted gene-level statistical association tests that involve mapping variants to genes and aggregating the variant-level associations using MAGMA.^28,38^ At a significance threshold of P-value<2.6 x 10^-6^ (accounting for testing 18,802 genes across the genome) we identified four genes (in 3 regions) and five genes (in 3 regions) in analysis with the Indian BMC GWAS (Figure 2a) and multi-ancestry meta-analysis (Figure 2b), respectively (Supplementary table 12 & 13). Majority of these were within regions identified by single variant analyses, with the exception of the well-known tumour suppressor gene *NOTCH1* (P-value=6.07×10^-7^), identified using multi-ancestry meta-analysis. Pathway analysis of genes with nominal significance (MAGMA P-value<0.05) identified two immune-related pathways interferon gamma signalling and PD-1 signalling; (Supplementary table 14) at P-value<0.05 after multiple comparisons adjustment. ^41^ Both of these associations were driven largely by genes in the MHC region and after excluding MHC genes the top pathway was the “highly calcium permeable nicotinic acetylcholine receptors” gene set (p=2.35×10^-3^, P_adjusted_=0.20; (supplementary table 14). Indeed, we noted that several nicotinic receptor encoding genes (*CHRNA3/4/5/6* and *CHRNB1/4*) showed weak but nominally significant (p<0.05) associations in the MAGMA analyses (Supplementary table 13).

**Figure 2:**
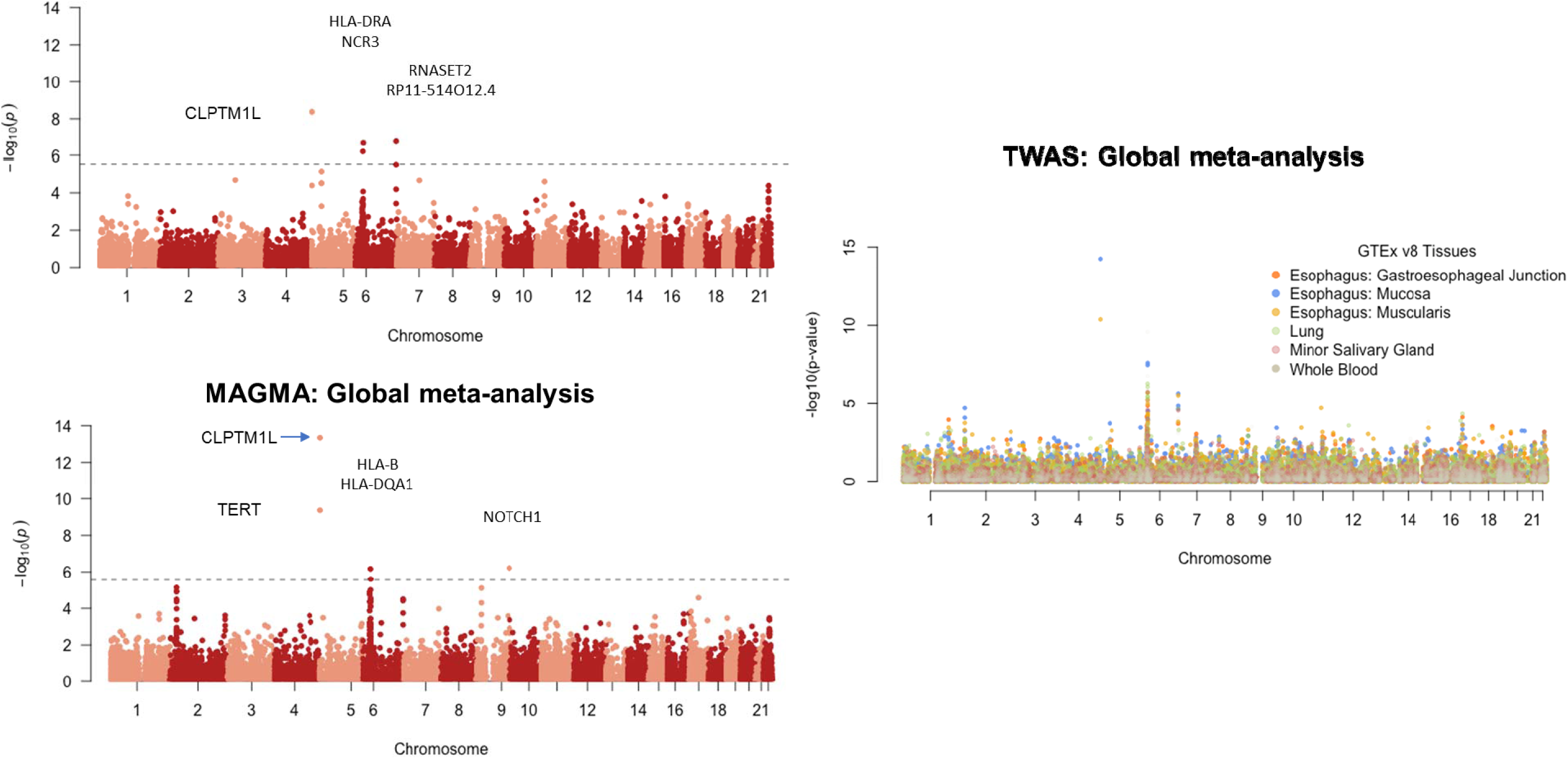
Genes associated with BMC. Manhattan plots for (A) gene-level statistical association tests using MAGMA (See **Methods**) for Indian BMC GWAS, (B) gene-level statistical association tests using MAGMA for multi-ancestry meta-analysis, (C) transcriptome-wide association analysis (TWAS) across 6 distinct tissues (See **Methods**) based on the multi-ancestry meta-analysis. In each plot, one dot represent the association of a gene with BMC.

We further carried out a transcriptome-wide association study (TWAS) using summary-statistics data from multi-ancestry meta-analysis and models for transcriptome prediction trained in GTEx across 6 tissues: 5 in upper gastrointestinal tract (Esophagus: gastroesophageal junction, Esophagus: Mucosa, Esophagus: Muscularis, Lung, Minor Salivary Gland) and Whole Blood. ^61^ In general, TWAS based on two types of esophagus tissues identified stronger signals for underlying genes in the known loci compared to other tissue (Figure 2c, Supplementary table 15). TWAS identified two genes *GPN1* (P-value=1.96×10^-5^) and *MS4A4A* (P-value=1.90×10^-5^) which were outside the known OC loci (Supplementary table 15). No signal for *GPN1* was evident in analysis restricted to Indian GWAS only (P-value=0.50; Supplementary table 15) and thus the finding seems to be primarily driven by the European GWAS. ^9^ In contrast, for *MS4A4A,* a gene known for its role in tumour immune microenvironments for several cancers, analysis restricted to India GWAS only showed nominal association (P-value=9.22 x 10^-4^) that manifested as a stronger association in TWAS using multi-ancestry meta-analysis, indicating potential shared mechanism across ancestries in this region (Supplementary table 15). ^62–64^

### Fine mapping and functional analysis of GWAS loci

To identify candidate causal variants underlying the associations identified, we performed fine mapping using Sum of Single Effects (SuSiE) for the genome-wide significant loci from the Indian BMC GWAS. ^48,49^ The analysis at 5p15.33 identified two potentially independent proximal signals with 28 and 9 variants in the 95% credible set, respectively (Figure 3 & Supplementary table 16) with the lead SNP of the locus rs31490 contained in the former credible set, mapping to the *CLPTM1L* gene. The latter credible set contained SNPs, mapping to the proximal *TERT* gene, that did not achieve genome-wide significance in the GWAS of Indian samples alone. However, this credible set included rs7713218 which was identified to be genome-wide significant in the multi-ancestry meta-analysis (Table 1), potentially tagging the conditionally independent association identified in the same region in analysis with Indian samples alone (Supplementary table 9). This highlighted the utility of the multi-ancestry analysis results in identifying shared effects across ancestries and that the 5p15.33 locus in particular, might contain two independent associations with BMC and other forms of Oral Cavity cancers. Fine mapping of the novel 6q27 locus produced a 95% credible set comprising of 17 variants (Figure 3, Supplementary table 16) while fine mapping in the 6p21.3 locus produced low-purity credible sets potentially due to the complicated LD structure of the HLA region (Supplementary table 16). To further characterize the functionality of SNPs significant at GWAS level, we performed a functional annotation (Supplementary table 17). ^28^ To nominate candidate genes which have strong evidence of transcriptomic association with BMC risk, we conducted summary based mendelian randomization (SMR) coupled with eQTL colocalization in blood, using summary data reported by the eQTLGen consortium (Supplementary table 18). ^39,40^ The SMR analysis leveraged the much larger sample size of the eQTL Gen Consortium (n=31,684) despite the lack of tissue specificity of this eQTL data said relative to the eQTL resources used in the TWAS reported above. We identified four genes in the SMR analysis and showed evidence of colocalization between the GWAS and eQTL associations (Supplementary Table 18) in gene *CHRFAM7A* and *GOLGA8R* at 15q13.2, *CSTF3* at 11p13, and *EP300* at 22q13.2 from the Indian data, while no significant evidence was found from the trans-ancestry meta-analysis. Notably, *CSTF3* and *EP300* were also associated in the MAGMA analyses with P-value=2.5 x 10^-5^ and P-value=3.3 x 10^-2^, respectively (Supplementary table 13).

**Figure 3:**
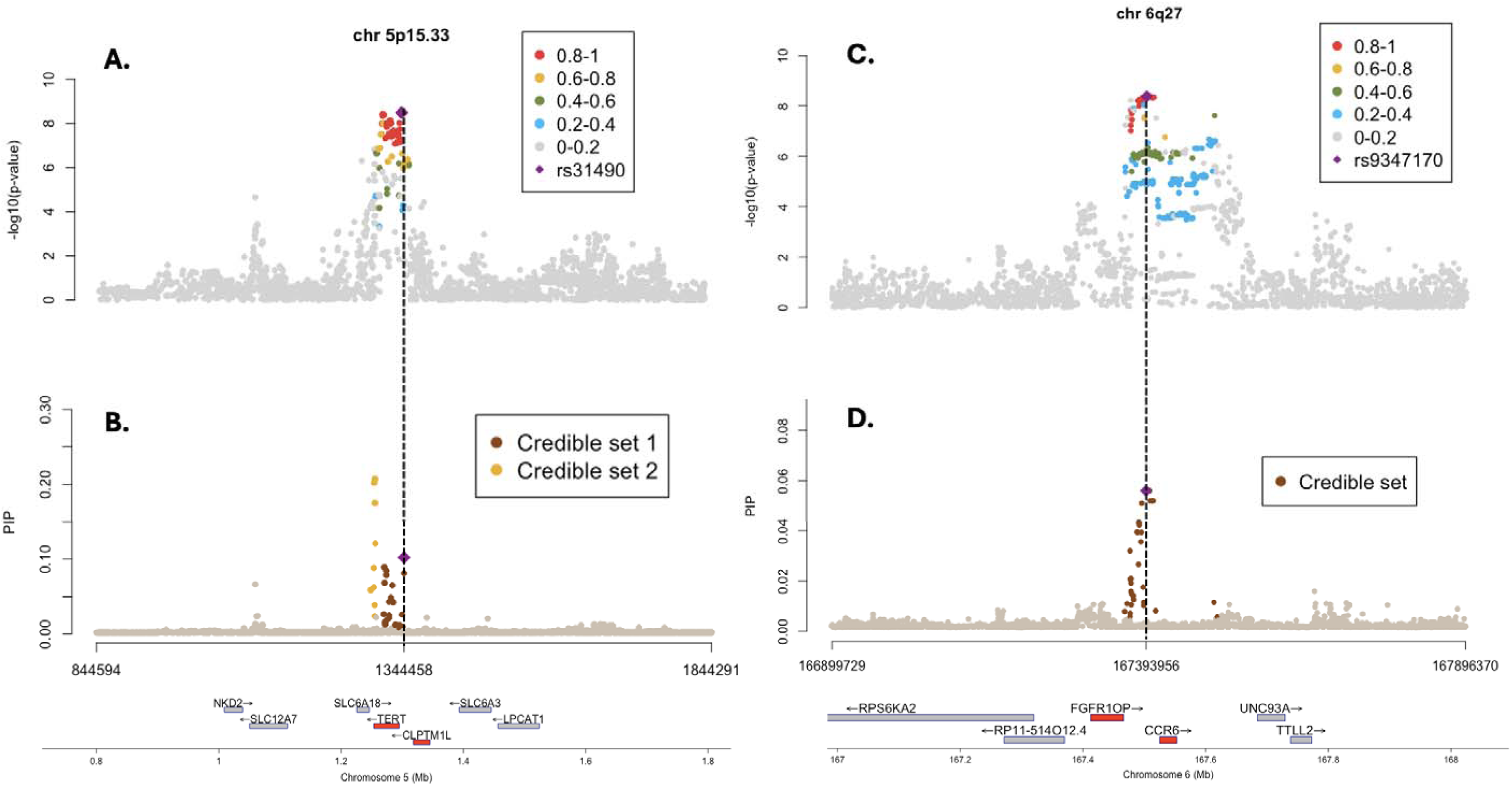
Fine mapping at Genome wide significant loci at 5p15.33 and 6q27. (A) Regional plot of 5p15.33 for with in rs31490 marked and other SNPs coloured according to LD (r2) with the index SNP in Indian samples. (B) Regional plot of ex SNP 5p15.33 showing the credible sets extracted by SuSiE (See **Methods**). SNPs are coloured according to their membership in each credible set. (C) Regional plot of 6q27 for with index SNP rs9347170 marked and other SNPs coloured according to LD (r^2^) with the index SNP in Indian samples. (D) Regional plot of 6q27 showing the credible set and SNPs are coloured according to their membership in credible set. For (A) and (C), the vertical axis represents the −log_10_(p-value) of the two-sided z-statistics, while for (B) and (D) it represents posterior inclusion probability estimated by SuSiE. For each plot the horizontal axis represents the genomic coordinates at the respective loci.

### Polygenic Analysis

We used the Genome-Wide Complex Trait Analysis (GCTA) tool to estimate the heritability of BMC cancer in India associated with common genotyped GWAS SNPs. ^37^ Assuming a 5-year prevalence of 0.26 per 1000 for oral cavity cancers, we estimated that 15.57% of BMC risk variation in the liability threshold scale could be explained by common SNPs included in our analysis (P-value=2.0 x 10^-11^). ^1^ We further estimated that the five lead SNPs identified in the Indian BMC GWAS explain 8.3% of the overall variation of BMC risk that can be explained by all the common SNPs together, in Indian population, and a corresponding polygenic score (PGS) could be expected to achieve an expected area under the curve (AUC) of 57.1% for predicting BMC risk in the Indian population. Further, we used POPCORN to estimate that cross-ancestry genetic correlation between BMC risk in India and OC risk in the EA population to be 0.53 (95% CI: 0.12-0.94), indicating significant overlap but also possible differences in genetic etiology for different forms of OC cancers (Supplementary figure 3). ^65^

Finally, we carried out a polygenic gene-environment interaction analysis using a genetic risk score based on the five lead SNPs which had genome wide significance in the Indian GWAS. We observed statistically significative evidence of multiplicative (P-value= 0.026) and additive (P-value=0.007) interactions between genetic risk score and tobacco chewing habit (Table 2). The pattern implied that the relative risk of BMC associated with higher level of the PGS was weaker for individuals who were at high risk due to chewing tobacco. At the same time, the risk differences associated with increasing value of PGS were stronger among tobacco chewers and *vice versa*. We estimate that the cumulative risk of developing BMC between age 40 and 74 for tobacco chewers was 0.6% and 0.3% in the highest and lowest genetic risk score categories respectively (Figure 4). The individuals with tobacco chewing habits and high genetic scores are expected to develop BMC ten years earlier compared to individuals with chewing habits and low genetic risk score.

**Figure 4:**
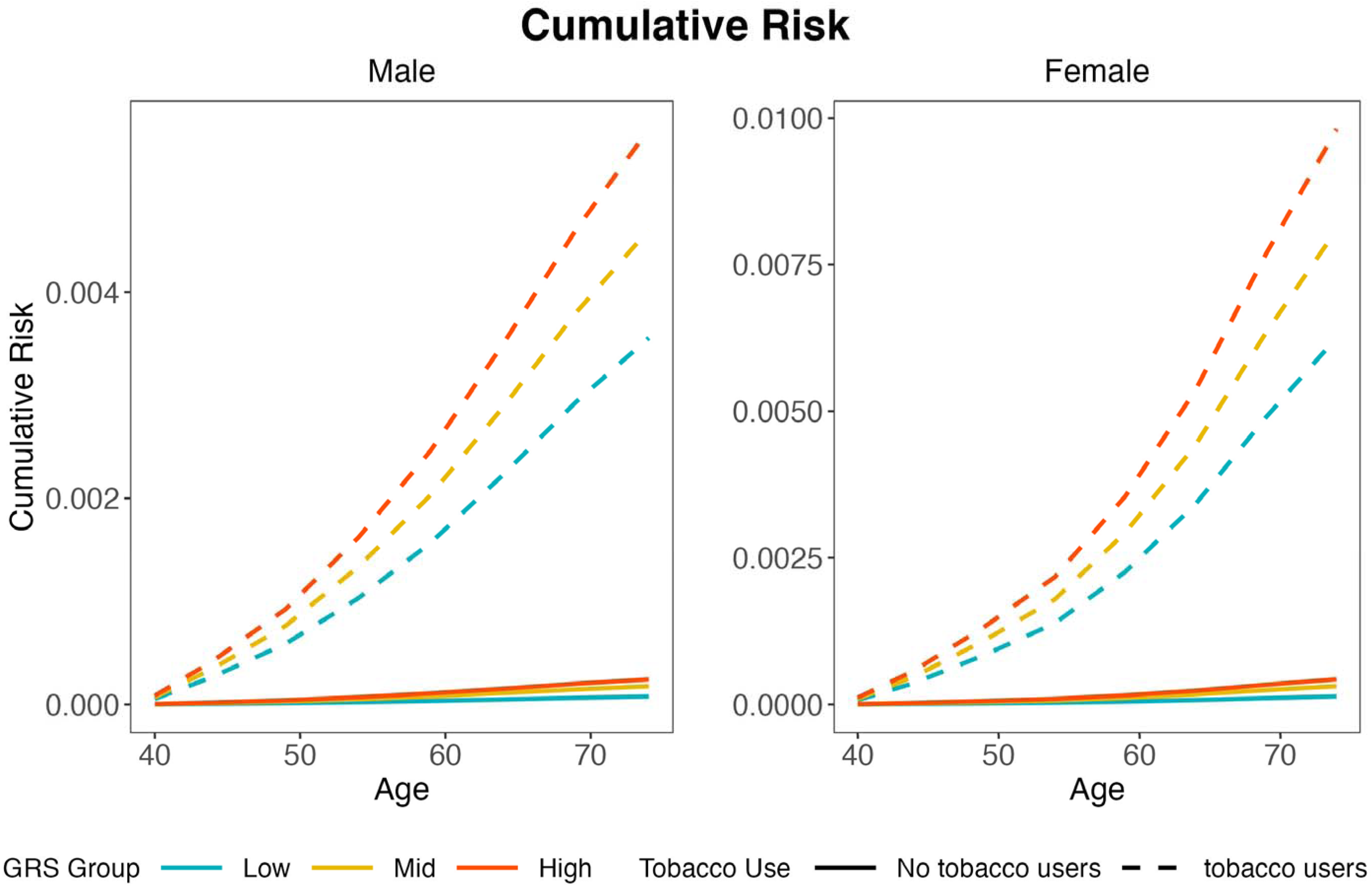
Risk stratified by genetic risk score and tobacco use. Cumulative risk (age 40-74) of developing Buccal Mucosa cancer among tobacco chewers with maximum and minimum genetic risk score stratified by gender.

**Table 2.** Joint effect between tobacco chewing and Indian BMC genetic risk score estimated on multiplicative scale.

| Indian BMC GRS score | Odds Ratio (95% Confidence Interval) |  |  |
| --- | --- | --- | --- |
|  | No Tobacco Users | Tobacco Chewers | Relative excess risk |
| <2 | Reference | 45.84 (28.91, 72.68) |  |
| 2-3 | 2.28 (1.39, 3.74) | 59.39 (37.71, 93.53) | 12.27 (0.62, 23.92) |
| 4-7 | 3.13 (1.75, 5.62) | 72.00 (43.48, 119.24) | 24.03 (2.15, 45.9) |
| Test for 1-df multiplicative interaction effect: P-value=0.026. |  |  |  |
| Test for 1-df multiplicative interaction effect using retrospective likelihood: P-value=0.007. |  |  |  |

## Discussion

We conducted the first GWAS of buccal mucosa cancer susceptibility in India which faces a large burden of the malignancy due to high prevalence of tobacco chewing. Using our own data, we identified one novel risk locus, 6q27 for BMC, and further using multi-ancestry meta-analysis of GWAS we identified one novel oral OC locus at 9q34.3 near the tumour suppressor gene *NOTCH1*, and a novel signal for OC risk within the 5p15.13 region near the gene *TERT*. We further confirm the role of multiple HLA loci for BMC susceptibility in India, indicating a potential role of the underlying immune component. Our heritability and co-heritability analyses indicate significant potential of common SNPs to explain variation of risk of BMC in the Indian population beyond the identified variants, and that there is both significant overlap and heterogeneity in the genetic architecture of BMC with other forms of OC. Finally, based on a polygenic analysis, we provide unique insight into the nature of gene-environment interactions in the etiology of BMC.

We identified a novel risk locus, 6q27, based on the Indian BMC GWAS. The lead SNP rs9347170 in this region is near the *CEP43* gene, and a recent proteomic based Mendelian Randomization study has identified the gene as a causal target for the risk of lung cancer. ^66^ The 6q27 locus also contains other tumour suppressor genes such as *DLL1* involved in *NOTCH1* cell signaling pathway. The effect allele(C) for rs9347170 is common across multiple global populations, but it did not show significant association with the risk of OC in the EA GWAS, despite its larger sample size (Table 1 & Supplementary table 8). In the cross-ancestry analysis, the locus was found to be genome-wide significant indexed by an alternative lead SNP rs204295, but underlying heterogeneity in effect sizes show that association at this locus is largely driven by the Indian BMC GWAS (Table 1 & Supplementary table 11). Further, the MAGMA analysis of cross-ancestry GWAS meta-analysis identified the major tumour suppressor gene *NOTCH1* itself as a potential novel risk locus for OC (Figure 2, Supplementary table 12 & 13). Thus, overall these two novel findings implicate a potential role *of NOTCH1* signalling pathway on the development of BMC and perhaps more broadly on OC, which complements the established somatic role of *NOTCH1* mutations in OC. ^67^

The genomic region 5p15.13 is known to harbour many cancer susceptibility variants with at least two possible candidate genes *CLPTM1L* and *TERT*. ^68^ The lead SNP rs31490 identified in the Indian data for this region is 756 bp upstream of TSS of *CLPTM1L.* ^52–54^ In conditional analysis (conditioned on lead SNP in locus, rs31490), we observed genome-wide significant association in an upstream region indexed by the lead SNP rs2735846 (4.2 kb from canonical TSS site of TERT gene; Table 1 & Supplementary table 9) within the Indian GWAS. ^52–54^ Multi-ancestry meta-analysis identified two independent genome-wide significant signals (Table 1 & Figure 1), near the *CLPTM1L* and *TERT* genes, respectively, within the 5p15.13 and fine-mapping analysis within the India data also indicated existence of two independent credible sets (Figure 3 and Supplementary table 16). In totality, the Indian study provided new evidence of complex allelic heterogeneity for risk of OC within the 5p13.13 region. GWAS SNPs on 5p15.33 region have been previously associated with telomerase length implying that the *TERT* may be the gene targeted by some risk variants. Germline *TERT* promotor mutations have been identified in familial melanoma as well as *TERT* somatic mutations in multiple tumours. ^57,58,69^ The studies conducted in Asian populations for lung cancer indicated that the presence of long telomerase, conferred by genetic polymorphisms foster the survival of lung epithelial cells and heightens their propensity to undergo malignant transformation. ^70^

Our study confirms that HLA loci confer risk of BMC in the Indian population as has been shown for risk of OC broadly in European populations. The HLA system has long been associated with infection, inflammation and autoimmunity. ^71^ The lead SNP found in the Indian study, rs9270532, is 2 kb upstream transcript variant for *HLA-DRB1*, which has been related to acute and chronic hepatitis B virus persistent infection. ^72^ Two other lead SNPs in *HLA,* rs28430392 and rs17427431, were closest to *HLA-DQB1.* The CADD score for SNP rs28430392 (upstream gene variant) and rs17427431 in the HLA region (upstream gene variant) was 6.29 and 8.71, respectively (Supplementary table 8). ^73^ The HLA regions observed for BMC Indian GWAS was located around *HLA-DQB1/HLA-DRB1* region while the SNPs identified in HLA region for oropharyngeal cancer were distributed in a wide range, covering multiple HLA genes including *HLA-B, HCP5, HLA-DRA1, HLA-DRB,1 HLA-DQA1 and HLA-DQB1* in European and North American population. ^7,9^ The association of SNPs in HLA region in European population were attributed to HPV positive OPCs. ^9^ When we did HPV sequencing on tumour samples (n=120) on the BMC cancer cases included in GWAS, none of them were found to be positive for HPV (data not published). Thus, while the association of HLA loci with BMC risk in India indicates an infectious/immune component for this cancer in the Indian population, the mechanism of risk may not solely be due to HPV infection. The involvement of a distinct immune component is also supported by the pathway analysis findings of PD-1 and interferon gamma signalling.

While a number of loci previously identified based on OC GWAS did show evidence of association in the Indian BMC GWAS, evidence lacked for two loci, 2p23.3 and 4q23.3. Of particular note is the locus 4q23.3 containing the gene *ADH1B,* an alcohol metabolizing gene that has been associated with many alcohol-related conditions including oral cancer based on EA GWAS. While the frequency of the effect allele for the index SNP at this locus was low in the Indian population (6%), we estimated the power of the Indian BMC GWAS for detecting the association at a nominal level and at the reported effect-size for the SNP in the EA GWAS to still high (>90%). Further, the frequency for the effect allele for the lead SNP at the locus 2p23.3 was quite high in the Indian population (47%) and thus the lack of replication at this locus also does not appear to be due to power. Genetic correlation analysis of the Indian BMC GWAS and EA OC GWAS further indicated that while there is substantial overlap in the underlying genetic architecture, there is also likely to be significant heterogeneity due to differences in the site of cancer, underlying environmental exposures, and other population characteristics. In summary, at the level of genetic correlation some SNPs and genetic loci were specific to each population putting BMC as a distinct biological entity. Heritability analysis indicates 15% of BMC risk variation in the Indian population can be explained by common genetic variations included in GWAS studies. The five identified variants (Table 1) from our study only explain a small fraction of this heritability and larger BMC GWAS in the future will help to identify many additional susceptibility loci, both specific to BMC and for OC more broadly.

BMC in India is considered largely driven by tobacco chewing, a habit that is highly prevalent in India (around 30%). ^74^ Within our own study, we observe BMC risk is increased among tobacco chewers by 28-fold (OR=27.85, 95%CI= 22.10-35.08), compared to non-tobacco users. We examined the interaction of chewing tobacco habit and genetic predisposition defined by the 5 identified SNPs for BMC risk based on the Indian study. Our polygenic analysis (Table 2) detected significant evidence of sub-multiplicative interactions, implying that the relative-risk associated with one factor in attenuated in the presence of the other factor. Nevertheless, we still found that, information on genetic predisposition can provide important stratification of absolute risk for tobacco chewers (Figure 4), and further discovery of genetic risk loci for BMC may be useful for risk stratification for the high-risk tobacco chewers. Overall, our study showcases the value of genetic association studies for conditions with significant lifestyle related risk factors, offering insight into the nature of gene-environment interactions and its implications for risk stratification.

While our study provides several novel insights into the genetic etiology of BMC, it has some limitations as well. First, the sample size for our current GWAS is modest, especially by the standard of the latest GWAS of many common diseases. As a result, the number of discoveries we have made are limited and the precision of estimates of heritability and co-heritability parameters are low. Clearly, larger studies for BMC indication oral cancers are needed in the future to continue the discovery effort. Second, as oral tissues are not represented in major expression quantitative trait studies like GTEx, we were able to conduct transcriptome-wide association studies (TWAS) only based on blood and multiple surrogate tissues in upper gastrointestinal tract, such as lung, minor salivary gland and esophageal locations. We showed that TWAS based on esophageal tissues typically show stronger association for OC risk even though the sample size for the underlying eQTL studies for these tissues were considerably lower compared to some alternative tissues like lung and blood (Figure 3). Thus, it appears that in the future generation of QTL for expression traits for oral tissues will be beneficial for the interpretation of findings from BMC and OC GWAS.

In conclusion, we conducted the first GWAS of BMC risk in India which faces a special burden of the malignancy due to high prevalence of tobacco chewing. Our study, together with a smaller GWAS of BMC from Taiwan and existing GWAS of OC from European ancestry populations, identifies novel risk loci and fine-mapped variants, and provides unique insight into the genetic architecture of BMC and its overlap with other forms of OC. Our study also provides an important perspective into the potential nature of polygenic gene-environment interactions in diseases that have major lifestyle related risk factors.

## Author contributions

SM and RD designed, conceptualized, and directed the study. SM, PC, AM, and MK monitored its progress. SM, PD, and SG supervised the laboratory activities, while SM, RD, NC, SK, and DD oversaw the data analysis processes. SM, DD, AI, SK, YD, ZW, and OJ conducted the formal analysis. The manuscript was written by SM, DD, AI, RD, NC, PR, PC, SK, AC, and SK, with GG providing editing assistance. All authors provided critical feedback and helped shape the research analysis and manuscript. AC and CW curated and edited the Taiwan data study.

## Data Availability

All data produced in the present study are available upon reasonable request to the authors and will be made publicly available upon publication of the manuscript in a peer-reviewed scientific journal.

## Acknowledgements for Taiwan Head and Neck Cancer Study (THANCS)

The authors thank Dr. Tseng-Cheng Chen and Dr. Pei-Jen Lou (National Taiwan University Hospital); Dr. Ming-shui Tsai and Dr. Chun-Hung Hua (China Medical University Hospital); Dr. Chung-Jan Kang (Chang Gung Memorial Hospital, Linkou); and Dr. Chi-Yen Chien (Chang Gung Memorial Hospital, Kaohsiung). The authors also thank the study staff and the study participants.

## Acknowledgements for Centre for cancer Epidemiology, Tata Memorial Centre

We would like to sincerely acknowledge all the participants, especially the patients, who actively provided their information. The staff at the Division of Molecular Epidemiology and Population Genomics, CCE, have relentlessly contributed to laying the foundations of this research work.

