## Supplementary figures and images for "A Genome-wide Association study of Buccal Mucosa Cancer in India and Multi-ancestry Meta-analysis Identifies Novel Risk Loci and Gene-environment Interactions"

### Supplementary Figure 1

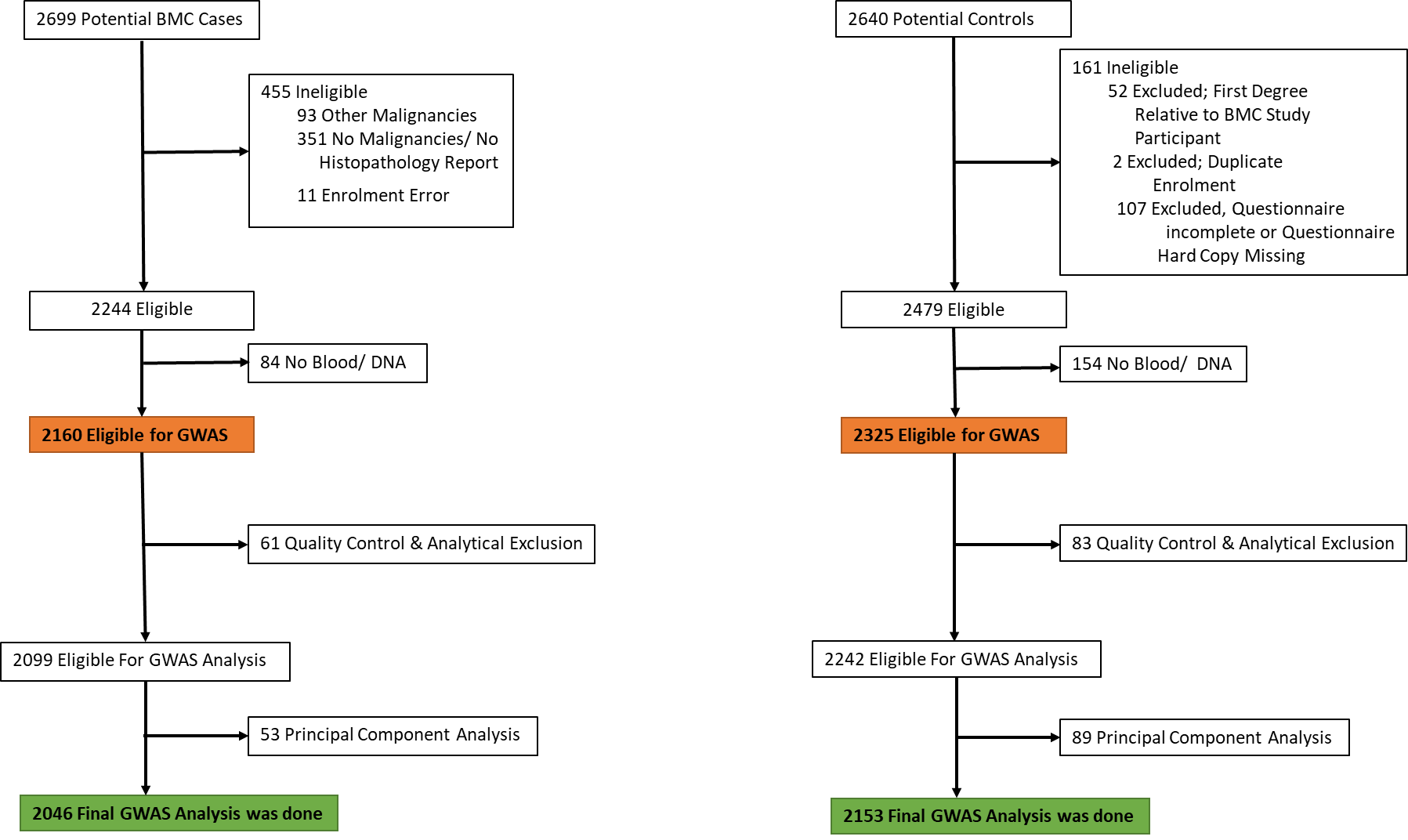

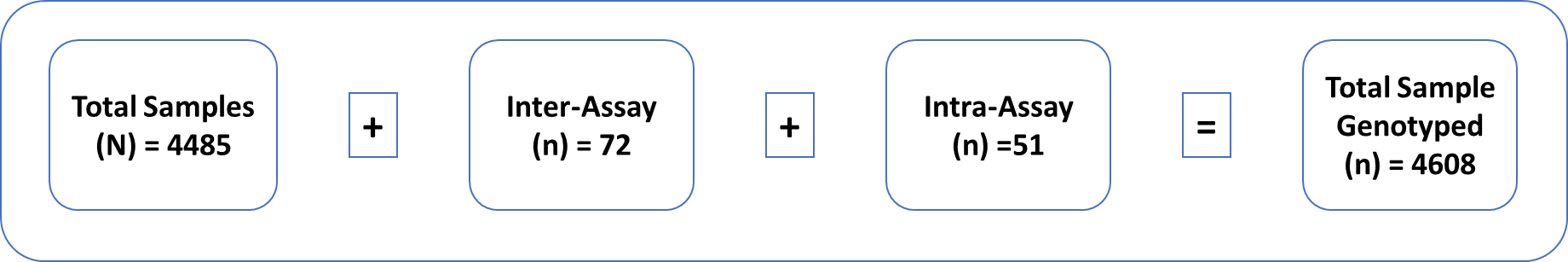

### Supplementary Figure 2

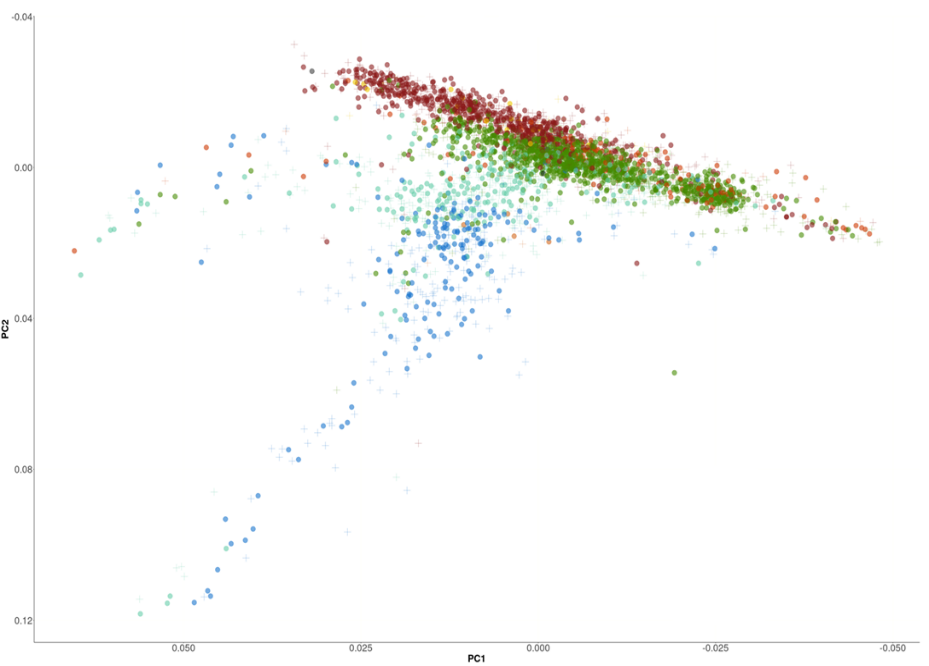

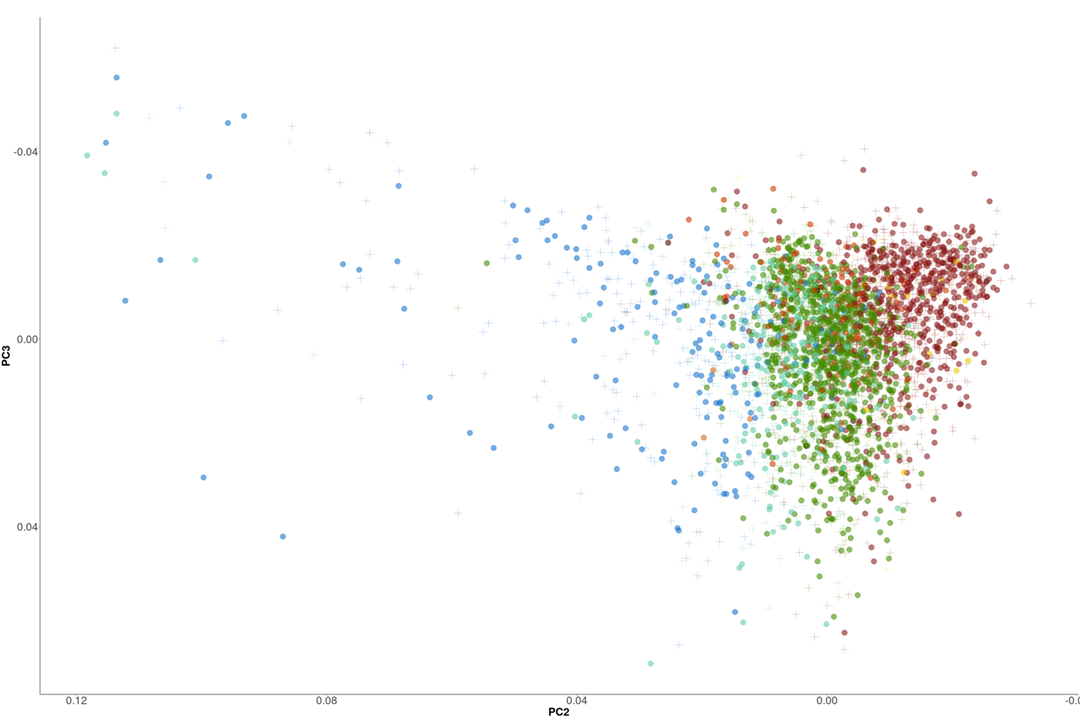

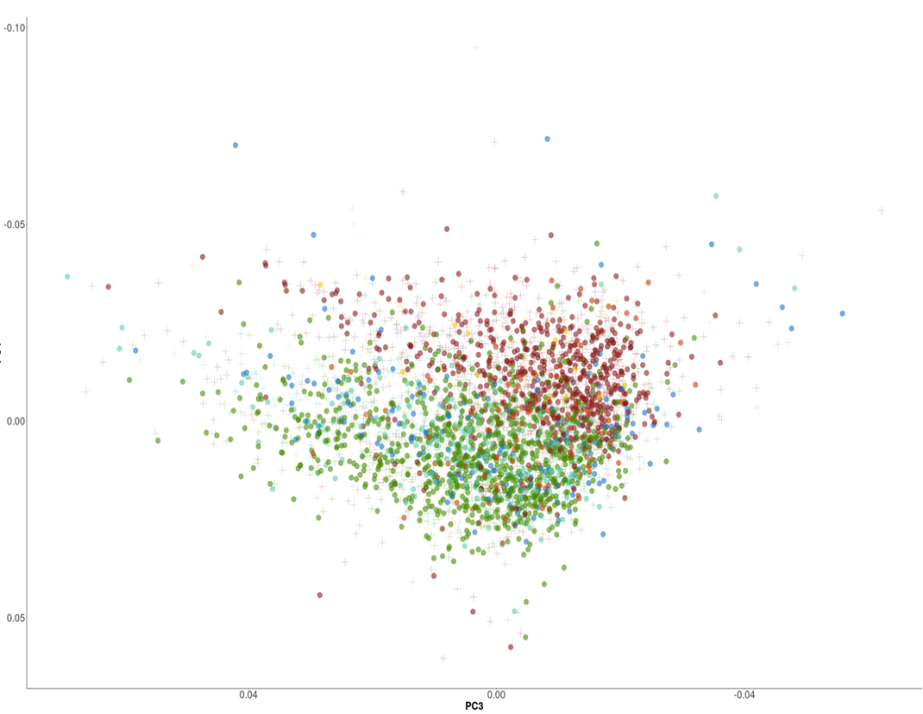

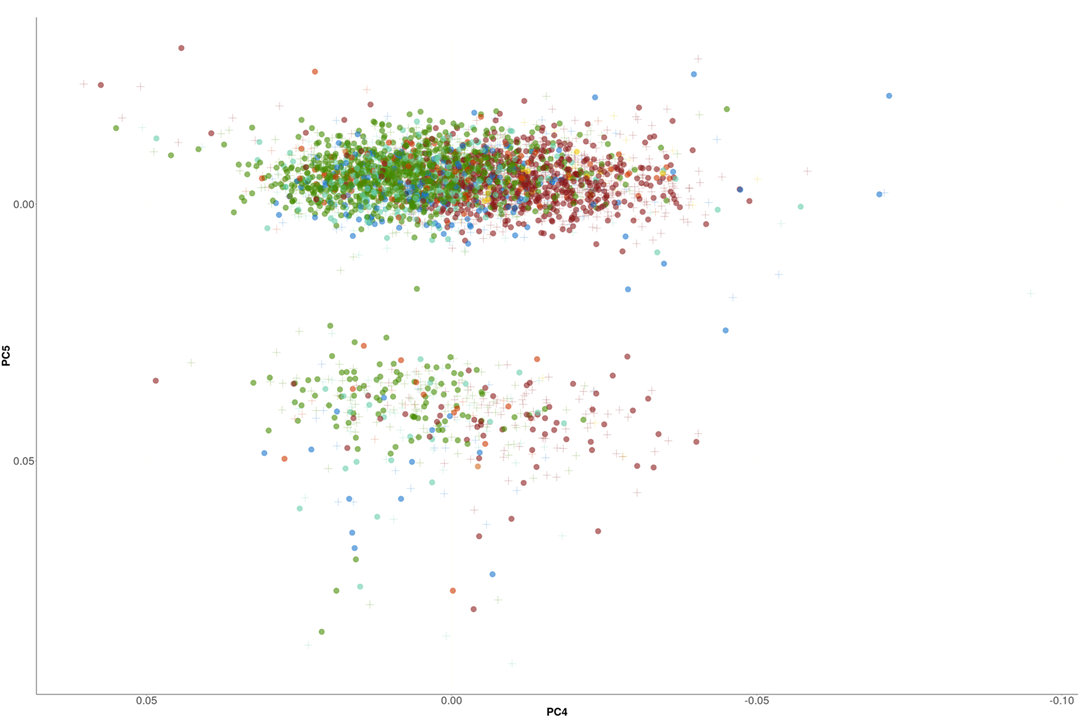

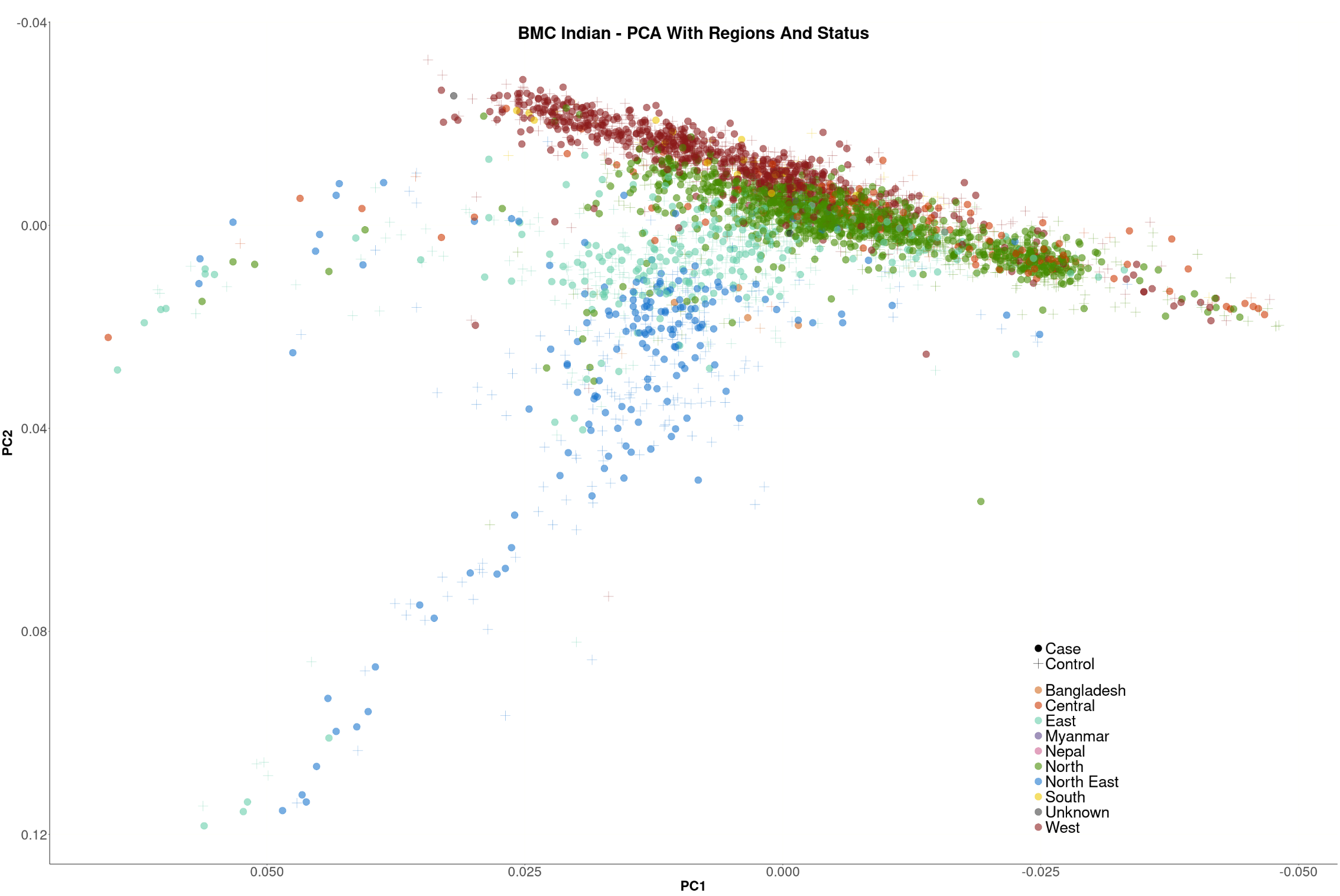
