## Supplementary Figure 3 for "A Genome-wide Association study of Buccal Mucosa Cancer in India and Multi-ancestry Meta-analysis Identifies Novel Risk Loci and Gene-environment Interactions"

**Supplemental Figure 3.** POPCORN analysis estimating genetic correlation across trans-ancestry populations regarding oral cavity cancer risk.

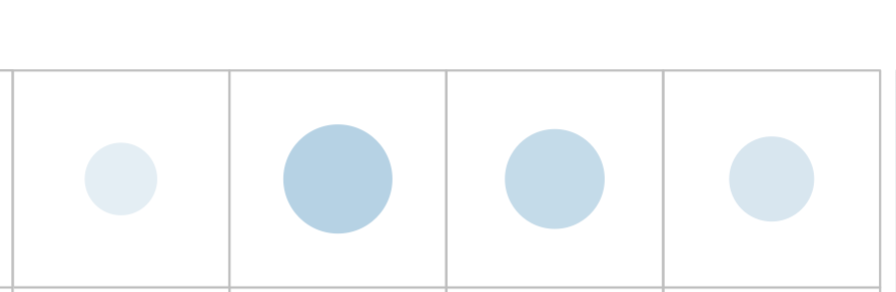

**Europe**

**Hispanic**

**Taiwan**

**North**

**America**

**India**

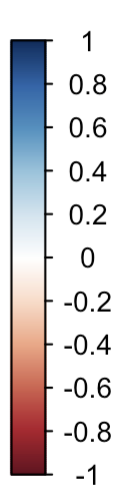
